# Transdiagnostic neurocognitive deficits in patients with type 2 diabetes mellitus, major depressive disorder, bipolar disorder, and schizophrenia: A 1-year follow-up study

**DOI:** 10.1101/2021.04.21.21255819

**Authors:** Patricia Correa-Ghisays, Joan Vicent Sánchez-Ortí, Vicent Balanzá-Martínez, Gabriel Selva-Vera, Joan Vila-Francés, Rafael Magdalena-Benedito, Victor M Victor, Irene Escribano Lopez, Antonio Hernández-Mijares, Juliana Vivas-Lalinde, Constanza San-Martín, Benedicto Crespo-Facorro, Rafael Tabarés-Seisdedos

## Abstract

**Background:** Impairments in neurocognition are critical factors in patients with major depressive disorder (MDD), bipolar disorder (BD), and schizophrenia (SZ), and also in those with somatic diseases such as type 2 diabetes mellitus (T2DM). Intriguingly, these severe mental illnesses are associated with an increased co-occurrence of diabetes (direct comorbidity). This study sought to investigate the neurocognition and social functioning across T2DM, MDD, BD, and SZ using a transdiagnostic and longitudinal approach.

**Methods:** A total of 165 subjects, including 30 with SZ, 42 with BD, 35 with MDD, 30 with T2DM, and 28 healthy controls (HC), were assessed twice at a 1-year interval using a comprehensive, integrated test battery on neuropsychological and social functioning.

**Results:** Common neurocognitive impairments in somatic and psychiatric disorders were identified, including deficits in short-term memory and cognitive reserve (p < 0.01; η^2^p = 0.08-0.31). Social functioning impairments were observed in almost all the disorders (p < 0.0001; η^2^p = 0.29-0.49). Transdiagnostic deficits remained stable across the 1-year follow-up (p < 0.001; η^2^p = 0.13-0.43) and could accurately differentiate individuals with somatic and psychiatric disorders (χ^2^ = 48.0, p < 0.0001).

**Conclusions:** This longitudinal study provides evidence of the overlap in neurocognition deficits across somatic and psychiatric diagnostic categories, such as T2DM, MDD, BD, and SZ, which have high comorbidity. This overlap may be a result of shared genetic and environmental etiological factors. The findings further lay forth promising avenues for research on transdiagnostic phenotypes of neurocognition in these disorders, in addition to their biological bases.

## INTRODUCTION

Type 2 diabetes mellitus (T2DM), major depressive disorder (MDD), bipolar disorder (BD), and schizophrenia (SZ) are complex and chronic illnesses with profound human and socioeconomic consequences.[1-2] Furthermore, these disorders co-exist more frequently than expected (direct comorbidity). For instance, the prevalence of T2DM in patients with MDD, BD, or SZ is 1.2–3-fold higher than that in the general population.[3]

Many studies have identified neurocognitive impairments in patients with MDD, BD, SZ,[4] and other chronic diseases, such as T2DM,[5] with serious consequences on daily functioning (e.g., conversing, driving, or walking). Unfortunately, on more occasions than desirable, the neurocognitive deficits remain unrecognized by physicians.[6] Furthermore, neurocognitive dysfunction in patients with T2DM is associated with poor self-management and an increased incidence of diabetes-related complications, including dementia in later life, presumably due to diabetes-related brain injuries.[7] Moreover, for older adults with type 1 diabetes mellitus (T1DM) or T2DM, depression significantly increased the risk of all-cause dementia.[8-9] While little or nothing is known on the effect of comorbidities on neurocognition and functional outcome measures in elderly patients with T2DM and BD or SZ, we may assume that they are at a greater risk of neurocognitive decline or dementia over time, since diabetes has been associated with more severe neurocognitive deficits in patients with SZ and BD.[10]

Given that there are direct comorbidities between these diseases and because neurocognitive dysfunctions are very common in patients with T2DM, MDD, BD, and SZ, we considered that neurocognitive alterations transgressed nosological boundaries. Identifying shared neurocognitive impairments may further clarify pathophysiological processes and perhaps reduce the impact of neurocognitive decline and dementia in older adults with these diseases. This study sought to investigate the following through a transdiagnostic and longitudinal approach: a) the neurocognitive profiles and functional outcome of patients with T2DM, MDD, BD, and SZ, compared to that in healthy controls (HC), over 1 year of follow-up; b) whether there are transdiagnostic deficits in terms of neurocognition underlying all somatic and psychiatric disorders; and c) the accuracy of the transdiagnostic deficits with regard to classifying patients into categorical diagnoses.

## MATERIALS AND METHODS

### Study design and ethical considerations

This article shows partial results of a more extensive study seeking to identify and validate peripheral biomarkers for neurocognitive deficits in MDD, BD, SZ, and T2DM. Only the variables that could advance the aim of this study were included in the analyses. This prospective, comparative cohort study was conducted between April 2015 and January 2018. Several biomarkers, clinical data, sociodemographic data, neurocognitive performance data, and social functioning data were collected at baseline (T1) and after 1 year (T2). Individuals with severe mental illness (SMI) were recruited from mental health units (MHU) of several towns in the province of Valencia, Spain (Foios, Catarroja, Paterna, and Sagunto), the psychiatry outpatient clinic and endocrinology department of the University Hospital Dr. Peset, and from the MHU of the Health Center of Miguel Servet, Valencia City, Spain. HCs consisted of residents of the same regions as the individuals with SMI. We compared them in terms of sex, age, and years of education to the extent possible. The study procedures were explained to participants, and all participants provided informed consent. The ethics committee or the institutional review board at each participating center approved the study protocol, and the study was conducted according to the ethical principles of the Declaration of Helsinki.

### Participants

At T1, the sample consisted of 165 subjects, including 30 individuals with SZ, 42 individuals with BD, 35 individuals with MDD, 30 individuals with T2DM, and 28 genetically unrelated HCs. At T2, there were 125 subjects, including 27 individuals with SZ, 29 individuals with BD, 25 individuals with MDD, 25 individuals with T2DM, and 19 HCs. SZ, BD, and MDD were diagnosed according to the criteria of the Diagnostic and Statistical Manual of Mental Disorders (DSM-5).[11] T2DM was diagnosed based on the Standards of Care criteria of the American Diabetes Association.[12] For recruitment as HCs, the absence of physical illness, pharmacological treatments, and family history of SMI in first-degree relatives was required. In addition to being diagnosed with one of the conditions mentioned above, the other inclusion criterion was an understanding and provision of written consent. For MDD and BD, it was necessary to meet the remission criteria of an acute affective episode, and individuals with SZ had to be clinically stable. Individuals with T2DM had to be free of severe diabetic neuropathy and kidney disease (serum creatinine <1.5 mg/dl). General exclusion criteria for all groups included: clinical conditions that hindered the study design, current hospitalization, documented neurocognitive impairment (intellectual disability or dementia), disability or inability that prevented an understanding of the protocol, current substance abuse disorder, pregnancy, intake of corticosteroids, antioxidants, antibiotics, and immunologic therapies, fever over 38°C, and history of vaccination within four weeks of the evaluation. The same inclusion and exclusion criteria were used at T1 and T2.

### Assessments

The assessments were conducted by the same experienced psychologists and psychiatrists of the research group. Sociodemographic data, including sex, age, and years of education, were collected at T1. For individuals with SMI, the age of disease onset, illness duration, episodes, admissions, psychopharmacological treatment, and total medication were assessed.

Clinical evaluations were conducted using the following scales: (i) the Hamilton Depression Rating Scale (HDRS),[13] (ii) the Young Mania Rating Scale (YMRS),[14] (iii) the Positive and Negative Symptoms Scale (PANSS),[15] which was also used to assess the severity of illness in psychiatric individuals, and (iv) the Clinical Global Impression scale (CGI).[16] The HDRS and YMRS were used for cases of BD and MDD that met the remission criteria (Euthymia = HDRS < 9 and YMRS < 7).

Social functioning was evaluated using: (i) the Functional Assessment Short Test (FAST),[17] (ii) the Short Form-36 Health Survey questionnaire (SF-36),[18] and (iii) the WHO Quality of Life-BREF (WHOQOL-BREF).[19]

Neurocognitive performance was evaluated using a battery of cognitive tests and subtests previously used by our group [20-24]. Test and subtests scores were divided into seven neurocognitive domains: 1) Learning and verbal memory (L&VM) [(i) Complutense Verbal Learning Test (TAVEC) V3, V8, and V10 variables,[25]] 2) Cognitive Flexibility (CF) [(ii) Stroop Color and Word test (SCWT) Color/Word subtest,[26] and (iii) Wisconsin Card Sorting Test (WCST) Categories Completed and Perseverative Errors scores,[27]] 3) Verbal Fluency (VF) [(iv) Verbal Fluency Tasks (VFT) Semantic and Phonemic forms,[28-29]] 4) Working memory (WM) [(v) Trial Making Test (TMT) Part B,[30] and (vi) Wechsler Adult Intelligence Scale 3rd version (WAIS-III) Digit Span-B subtest,[31]] 5) Short-term Memory (StM) [TAVEC V1 and V4 variables,[25] and WAIS-III Digit Span-A subtest,[31]] 6) Visual Memory (VM) [(vii) Rey-Osterrieth Complex Figure Test (ROCFT),[32]] 7) Processing Speed (PS) [(viii) Finger Tapping Test (FTT),[30, 33] WAIS-III Digit Symbol Coding subtest,[31] SCWT Color and Word subtests,[26] and TMT Part A,[30]] and three neurocognitive indices, including the Global Cognitive Score (GCS), which was calculated by averaging the seven neurocognitive domain scores; the premorbid Intelligence Quotient (IQ), which was calculated using the WAIS-III vocabulary subtest, considered a classical measure of the level of intelligence prior to the onset of a mental disorder [34]; and the Cognitive Reserve (CR), which was estimated based on the results of the WAIS-III Vocabulary subtest and the number of years of formal education.[35] **Figure 1** represents a summary of the instruments used to assess the clinical status, social functioning, and neurocognitive performance.

**Figure 1.**
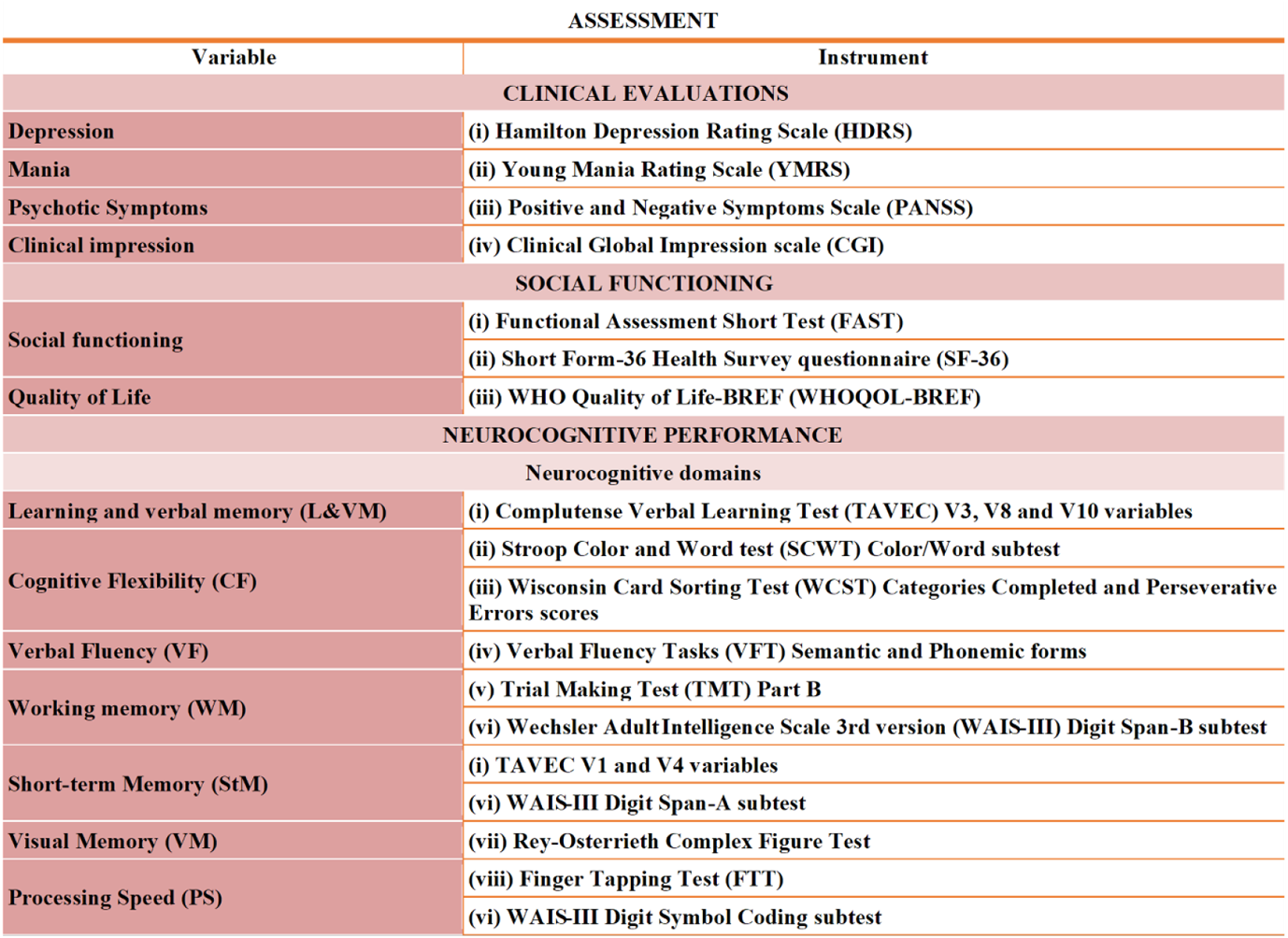
Summary of the tools used to assess the clinical status, social functioning, and neurocognitive performance.

### Statistical analyses

All statistical analyses were performed in R (version 3.3.1).[36] The direct scores obtained for neurocognitive and social functioning measures were transformed into Z-scores. For the calculation of the Z-scores, the mean and standard deviation of the HCs at T1 were taken as reference values. Analyses were conducted using a one-way analysis of variance (ANOVA), ANOVA-corrected, asymptotic general independence test for continuous variables, a negative binomial generalized linear model for discrete variables, and a Fisher’s Exact Test for categorical variables. Normality was assumed for all continuous variables because the sample is sufficiently representative of the target population, which was statistically verified. The between-group differences for the neurocognitive and social variables at T1 and T2 were assessed using a one-way analysis of covariance, with sex and age as covariates. A post-hoc analysis with a Bonferroni corrected pairwise t-test and Mann–Whitney U tests were performed to assess the between-group differences. To test the ability to discriminate between individuals with T2DM and SMI and HCs in terms of transdiagnostic neurocognitive deficits, a discriminant analysis and XGBoost linear regression were performed using a predictive model that included only the significant transdiagnostic neurocognitive deficits. For all analyses, p < 0.05 was considered statistically significant.

## RESULTS

### Sample description

A summary of the sociodemographic and clinical characteristics of the participants can be found in **Table 1**. Females represented about half of the total sample (47%). The mean age of the whole sample was 46.4 years, and the mean number of years of education was 12.5. The years of education were similar among the clinical groups. Depressive symptoms were more pronounced in the MDD group, and maniac symptoms were more prevalent in the BD and SZ groups. Individuals with SZ showed the highest scores in terms of psychotic symptomatology and the worst clinical impression. MDD and T2DM groups showed fewer episodes, admissions, intake of psychopharmacological treatment, and years of illness. The T2DM group had the highest age of onset.

**Table 1.** Sociodemographic and clinical characteristics of the sample at T1.

|  |  |  |  |  |  |  |  |
| --- | --- | --- | --- | --- | --- | --- | --- |
| **Variables<sup>a</sup>** | **HC** | **T2DM** | **MDD** | **BD** | **SZ** | **Statistical analyses** |  |
|  | (n=28) | (n=30) | (n=35) | (n=42) | (n=30) | **F(p)<sup>g</sup>** | **Post hoc test<sup>i</sup>** |
| *Sociodemographic* |  |  |  |  |  |  |  |
| **Sex<sup>b,h,j</sup>** | 18(64%) | 9(30%) | 24(68%) | 21(50%) | 7(23%) | 20.1**** | SZ,T2DM<HC,MDD<br>SZ<BD |
| **Age** | 36.6(14.5) | 57.3(9.3) | 47.3(11.8) | 50.0(9.5) | 40.8(10.7) | 15.3**** | HC<MDD,BD,T2DM<br>SZ,MDD<T2DM<br>SZ<BD |
| **Years of Education** | 16.1(3.3) | 12.5(5.8) | 11.9(4.3) | 11.6(4.4) | 10.4(3.3) | 7.1**** | SZ,BD,MDD,T2DM<HC |
| *Clinical* |  |  |  |  |  |  |  |
| **HDRS<sup>c</sup>** | 2.0(1.8) | 3.9(3.9) | 11.6(8.3) | 6.4(4.4) | 7.0(5.8) | 14.2**** | HC<BD,SZ,MDD<br>T2DM,BD,SZ<MDD |
| **YMRS<sup>c</sup>** | 0.8(1.6) | 1.5(2.2) | 1.9(2.6) | 3.5(4.5) | 3.2(4.9) | 3.4** | HC<BD |
| **PANSS-P<sup>c</sup>** | 7.0(0.0) | 7.0(0.0) | 7.0(0.3) | 8.5(3.8) | 10.6(4.3) | 10.6**** | HC,T2DM,MDD,BD<SZ |
| **PANSS-N<sup>c</sup>** | 7.0(0.0) | 7.1(0.7) | 8.4(4.9) | 10.3(6.5) | 18.6(10.1) | 20.1**** | HC,T2DM,MDD,BD<SZ |
| **PANSS-G<sup>c</sup>** | 16.0(0.0) | 17.0(2.3) | 19.8(8.6) | 22.7(9.9) | 31.8(12.7) | 16.9**** | HC,T2DM,MDD,BD<SZ<br>HC<BD |
| **CGI<sup>c</sup>** | - | 1.9(1.0) | 3.3(1.2) | 3.5(0.7) | 4.5(1.0) | 31.3**** | T2DM<MDD,BD,SZ<br>MDD,BD<SZ |
| **Age of onset<sup>d</sup>** | - | 44.3(9.8) | 35.3(12.1) | 26.5(8.6) | 25.6(8.0) | 25.6**** | SZ,BD,MDD<T2DM<br>SZ,BD<MDD |
| **Illness duration<sup>d</sup>** | - | 13.0(9.0) | 12.0(11.6) | 23.4(11.5) | 15.2(8.4) | 9.6**** | MDD,T2DM,SZ<BD |
| **Total episodes<sup>e</sup>** | - | - | 2.3(1.9) | 8.9(6.9) | 6.5(6.9) | 12.1**** | MDD<SZ,BD |
| **Total admissions<sup>f</sup>** | - | - | 0.1(0.6) | 1.4(1.9) | 1.7(1.6) | 10.4**** | MDD<BD,SZ |
| **Psychopharma-treatment** | - | 0.5(1.0) | 2.7(2.0) | 3.6(1.8) | 3.0(1.9) | 19.8**** | T2DM<MDD,SZ,BD |

<sup>a</sup> Expressed as mean(standard deviation) except when indicated, <sup>b</sup> female n(%). <sup>c</sup> Lower scores represent a better outcome. <sup>d</sup> Years. <sup>e</sup> Psychotic and mood episodes. <sup>f</sup> mental health unit. <sup>g</sup> ANOVA. <sup>h</sup> Chi-squared test. <sup>i</sup> Bonferroni test. <sup>j</sup> Mann-Whitney U test. Abbreviations: HC = Healthy Control, T2DM = Type-2 Diabetes Mellitus, MDD = Major Depressive Disorder, BD = Bipolar Disorder, SZ = Schizophrenia, HDRS = Hamilton Rating Scale for Depression, YMRS = Young Mania Rating Scale, PANSS = Positive and Negative Syndrome Scale, P = Positive, N = Negative, G = General, CGI = Clinical Global Impression, NS = Not Significant. (NS = $p > 0.05$ ; \* $p \leq 0.05$ ; \*\* $p \leq 0.01$ ; \*\*\* $p \leq 0.001$ ; \*\*\*\* $p \leq 0.0001$ ).

### Comparison of neurocognitive performance and social functioning between the groups

Neurocognitive performance and social functioning for all groups at T1 and T2 and between-group comparisons are shown in **Table 2**. With regard to neurocognitive performance by clinical groups, compared to that in HCs, the SZ group presented with significant differences in all ten variables at T2 and T1, except for IQ. The BD group presented with significant differences in seven variables at T1, except for VF, WM, and IQ. At T2, the BD group obtained similar results, except for IQ, which did not present significant differences. The MDD group presented significant differences at T1 in CF, StM, VM, PS, and CR, and in GCS at T2. The T2DM group presented significant differences at T1 in CR, and at T2 in StM and CR. With regard to social functioning in the clinical groups, compared to that in HCs, the SZ and BD groups presented significant differences in all three variables at T1 and only in SF-36 at T2. The MDD group presented with significant differences in all three variables at T1 and T2. In terms of social functioning, the T2DM group did not differ significantly from the control group. P-values were p < 0.0001, and significant effect sizes ranged from moderate-to-large at both time points (η^2^p = 0.08–0.50) in all previously mentioned situations. With regard to clinical group performance, deficits were more accentuated in the SZ and BD groups, followed by the MDD and T2DM groups, which showed milder to no deficits in some aspects of neurocognitive performance and social functioning. The MDD group showed poorer social functioning at both times **(Figure 2)**, with a large effect size (p < 0.0001; η^2^p = 0.29–0.31). The neurocognitive deficits shared by all clinical groups were StM and CR performance **(Figure 3)**.

**Table 2.** Outcomes at T1 and T2, and Between-group comparison (Z-scores)

| Variables<sup>a</sup> | HC |  | T2DM |  | MDD |  | BD |  | SZ |  | Statistical analyses |  |  |  |  |  |
| --- | --- | --- | --- | --- | --- | --- | --- | --- | --- | --- | --- | --- | --- | --- | --- | --- |
|  | T1<br>(n=28) | T2<br>(n=19) | T1<br>(n=30) | T2<br>(n=25) | T1<br>(n=35) | T2<br>(n=25) | T1<br>(n=42) | T2<br>(n=29) | T1<br>(n=30) | T2<br>(n=27) | T1<br>F(*p*)<sup>c</sup> | Post hoc test<sup>e</sup> | η<sup>2</sup>*p*<sup>g</sup> | T2<br>F(*p*)<sup>c</sup> | Post hoc test<sup>e</sup> | η<sup>2</sup>*p*<sup>g</sup> |
| Cognitive domains |  |  |  |  |  |  |  |  |  |  |  |  |  |  |  |  |
| L&VM | 0.0(0.9) | 0.5(0.9) | -0.9(1.0) | -0.3(1.1) | -0.6(1.3) | -0.4(1.3) | -1.3(1.3) | -0.8(1.2) | -1.8(1.2) | -1.7(1.2) | 8.6**** | SZ,BD<HC<br>SZ<T2DM,MDD | .18 | 9.3**** | SZ<MDD,T2DM,HC | .24 |
| CF | 0.0(0.7) | 0.2(0.8) | -1.0(1.1) | -1.2(1.2) | -1.0(1.3) | -0.7(1.3) | -1.4(1.2) | -1.7(1.4) | -1.9(1.5) | -1.7(1.5) | 9.4**** | SZ<BD,MDD,T2DM,HC | .19 | 6.8**** | SZ,BD<HC<br>SZ<T2DM,MDD | .18 |
| VF | 0.0(0.8) | 0.2(0.9) | -0.4(1.0) | -0.3(0.9) | -0.3(0.9) | -0.2(0.9) | -0.5(0.9) | -0.4(0.8) | -1.2(0.7) | -1.0(0.7) | 7.6**** | SZ<BD,T2DM,MDD,HC | .16 | 6.0**** | SZ<T2DM,MDD,HC | .17 |
| WM | 0.0(0.8) | 0.1(0.7) | -1.3(1.5) | -1.4(1.5) | -0.8(1.2) | -1.1(1.2) | -2.2(2.6) | -1.3(1.9) | -2.4(1.8) | -2.3(2.0) | 9.7**** | SZ,BD<T2DM,MDD,HC | .19 | 7.7**** | SZ<BD,T2DM,MDD,HC | .20 |
| StM | 0.0(0.6) | 0.6(0.8) | -0.7(0.6) | -0.6(0.6) | -0.6(0.7) | -0.6(0.8) | -0.8(0.7) | -0.7(0.6) | -1.4(0.7) | -1.2(0.7) | 13.1**** | SZ,BD,MDD<HC<br>SZ<BD,T2DM,MDD | .25 | 15.7**** | SZ,BD,T2DM,MDD<HC<br>SZ<BD,T2DM,MDD | .34 |
| VM | 0.0(1.0) | 0.6(0.8) | -1.1(1.7) | -0.6(1.6) | -1.3(1.7) | -1.2(1.7) | -2.2(1.4) | -1.5(1.5) | -1.6(1.3) | -1.5(1.3) | 8.5**** | BD,SZ<HC<br>BD<T2DM | .17 | 8.0**** | BD,SZ,MDD<HC<br>SZ<T2DM | .21 |
| PS | 0.0(0.6) | 0.2(0.5) | -1.1(1.1) | -1.2(1.0) | -1.1(1.0) | -1.2(0.9) | -1.7(1.3) | -1.4(1.0) | -1.9(1.2) | -1.9(1.2) | 15.8**** | SZ,BD<T2DM,HC<br>SZ<MDD | .28 | 14.0**** | SZ,BD,MDD<HC<br>SZ<BD,MDD,T2DM | .32 |
| Cognitive performance |  |  |  |  |  |  |  |  |  |  |  |  |  |  |  |  |
| GCS | 0.0(0.5) | 0.3(0.5) | -0.9(0.9) | -0.8(0.9) | -0.8(0.9) | -0.8(1.0) | -1.4(1.1) | -1.1(1.0) | -1.7(1.0) | -1.6(0.9) | 15.5**** | SZ,BD<T2DM,HC<br>SZ<MDD | .28 | 13.7**** | SZ,BD,MDD<HC<br>SZ<BD,MDD,T2DM | .31 |
| IQ | 0.0(1.0) | 1.5(1.0) | -0.2(1.2) | 0.7(1.2) | 0.0(1.3) | 1.0(1.1) | 0.0(1.3) | 0.1(1.4) | -1.0(1.5) | 0.1(1.1) | 3.4** | SZ<T2DM,MDD,BD | .08 | 4.7*** | BD,SZ<HC | .13 |
| CR<sup>b,d,f,h</sup> | 22(79%) | 17(90%) | 14(47%) | 14(56%) | 19(55%) | 13(52%) | 19(46%) | 11(38%) | 8(27%) | 7(26%) | 16.4** | SZ,BD,MDD,T2DM<HC<br>SZ<MDD | .31 | 20.1**** | SZ,BD,MDD,T2DM<HC<br>SZ<MDD,T2DM | .40 |
| Social functioning - Quality of life |  |  |  |  |  |  |  |  |  |  |  |  |  |  |  |  |
| FAST | 0.0(1.0) | 0.0(0.8) | -1.2(1.8) | -1.0(1.5) | -3.7(2.6) | -3.5(2.5) | -4.3(1.9) | -3.9(1.9) | -5.6(2.4) | -4.7(2.5) | 39.2**** | SZ<BD,MDD,T2DM,HC<br>BD,MDD<T2DM,HC | .49 | 22.7**** | SZ,BD,MDD<T2DM<br>SZ,BD,MDD<HC | .43 |
| SF-36 | 0.0(1.0) | 0.2(0.5) | -1.2(1.8) | -1.0(1.7) | -3.9(2.1) | -3.8(2.7) | -2.6(1.9) | -2.6(1.8) | -1.9(1.9) | -2.3(1.9) | 18.2**** | MDD<BD,SZ,T2DM,HC<br>BD,SZ<HC | .31 | 13.9**** | MDD<BD,SZ,T2DM,HC | .32 |
| WQB | 0.0(1.0) | 0.3(1.1) | -0.6(1.2) | -0.5(1.4) | -2.2(1.2) | -2.2(1.8) | -1.6(1.2) | -1.9(1.1) | -1.3(1.0) | -1.2(1.2) | 16.8**** | MDD<SZ,T2DM,HC<br>BD,SZ<HC | .29 | 12.2**** | MDD<SZ,T2DM,HC | .29 |

<sup>a</sup> Expressed as mean(standard deviation), <sup>b</sup> high n(%), <sup>c</sup> ANCOVA. <sup>d</sup> Chi-squared test. <sup>e</sup> Bonferroni test. <sup>f</sup> Mann-Whitney U test. <sup>g</sup> Partial Eta-Squared ( $\eta^2p$ ). <sup>h</sup> Correlation coefficient $\phi$ . Abbreviations: HC = Healthy Control, T2DM = Diabetes Mellitus Type 2, MDD = Mayor Depressive Disorder, BD = Bipolar Disorder, SZ = Schizophrenia, T1 = Time 1, T2 = Time 2, L&VM = Learning and Verbal Memory, CF = Cognitive Flexibility, VF = Verbal Fluency, WM = Working Memory, StM = Short-Term Memory, VM = Visual Memory, PS = Processing Speed, GCS = Global Cognitive Score, IQ = Intelligence Quotient, CR = Cognitive Reserve, WQB = WHO-QoL-BREF, NS = Not Significant. (NS = $p > 0.05$ ; \* $p \leq 0.05$ ; \*\* $p \leq 0.01$ ; \*\*\* $p \leq 0.001$ ; \*\*\*\* $p \leq 0.0001$ ). Effect size ( $\eta^2p$ : small $\approx 0.02$ ; moderate $\approx 0.15$ ; large $\approx 0.35$ . $\phi$ : small $\approx 0.10$ ; moderate $\approx 0.30$ ; large $\approx 0.50$ ).

**Figure 2.**
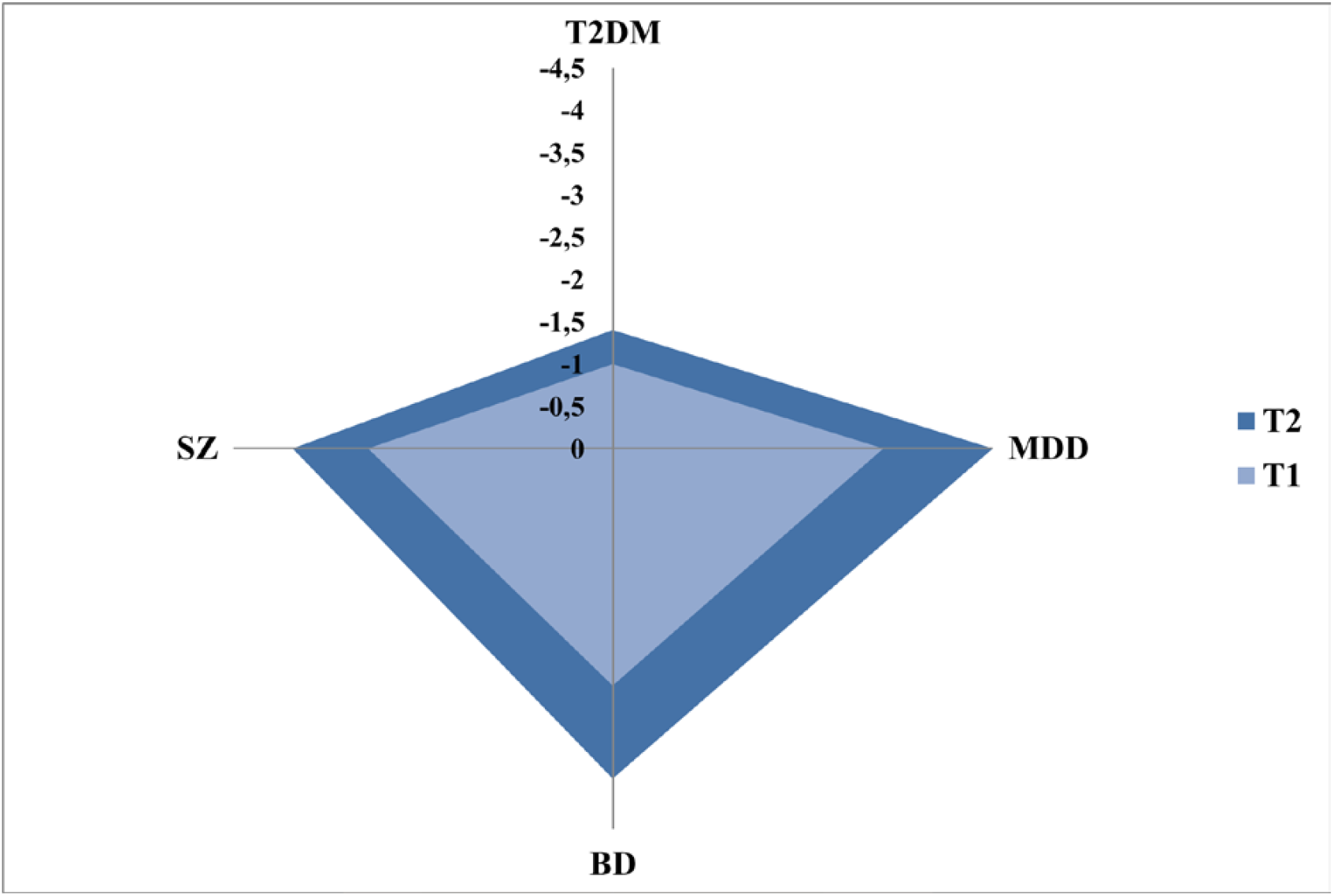
Degree of impairment of social functioning in the clinical groups. T2DM: Type-2 diabetes mellitus, MDD: Major depressive disorder, BD: Bipolar disorder, SZ: Schizophrenia, T1: Baseline data, T2: After one year

**Figure 3.**
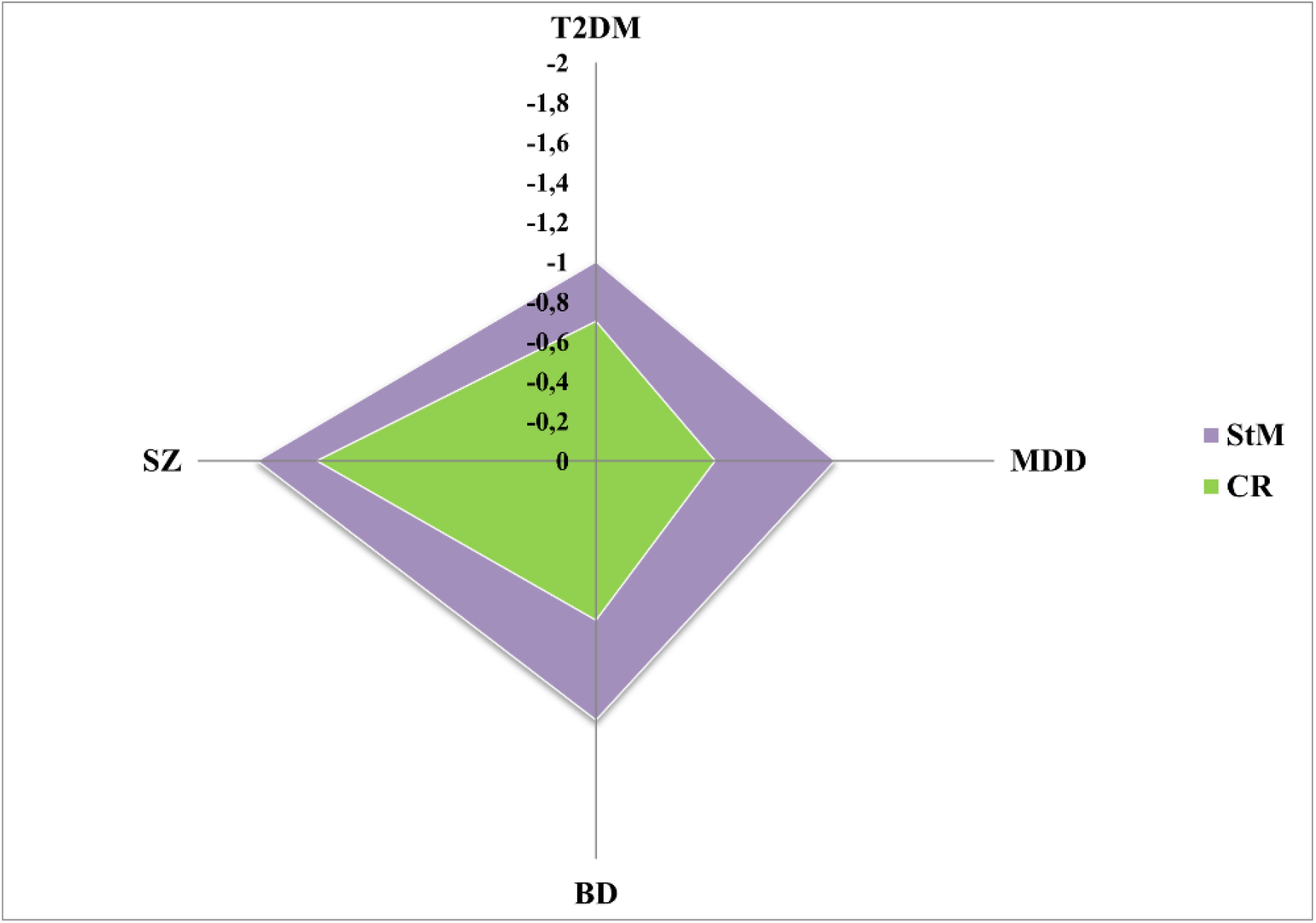
Shared transdiagnostic neurocognitive deficits by the clinical groups. T2DM: Type-2 diabetes mellitus, MDD: Major depressive disorder, BD: Bipolar disorder, SZ: Schizophrenia, StM: Short-term memory, CR: Cognitive reserve

### Significant transdiagnostic neurocognitive deficits between individuals with T2DM and SMI, and HC discriminate power

A stepwise discriminant analysis showed that for the significant neurocognitive transdiagnostic deficits included in this study, the best classified individuals with T2DM and SMI compared to HCs consisted of StM and both components of CR (years of education and IQ) (χ^2^ = 48.0, p < 0.0001). The XGBoost linear regression model revealed that both StM and CR have a strong capacity to discriminate between individuals with T2DM and SMI and HCs, with a correct classification rate of 78.3%. According to the model, when no other diagnostic information is available, storing, maintaining, and retrieving a certain amount of information for a few seconds, together with an adequate neurocognitive background, allows for the differentiation of individuals with T2DM and SMI from HCs **(Figure 4)**.

**Figure 4.**
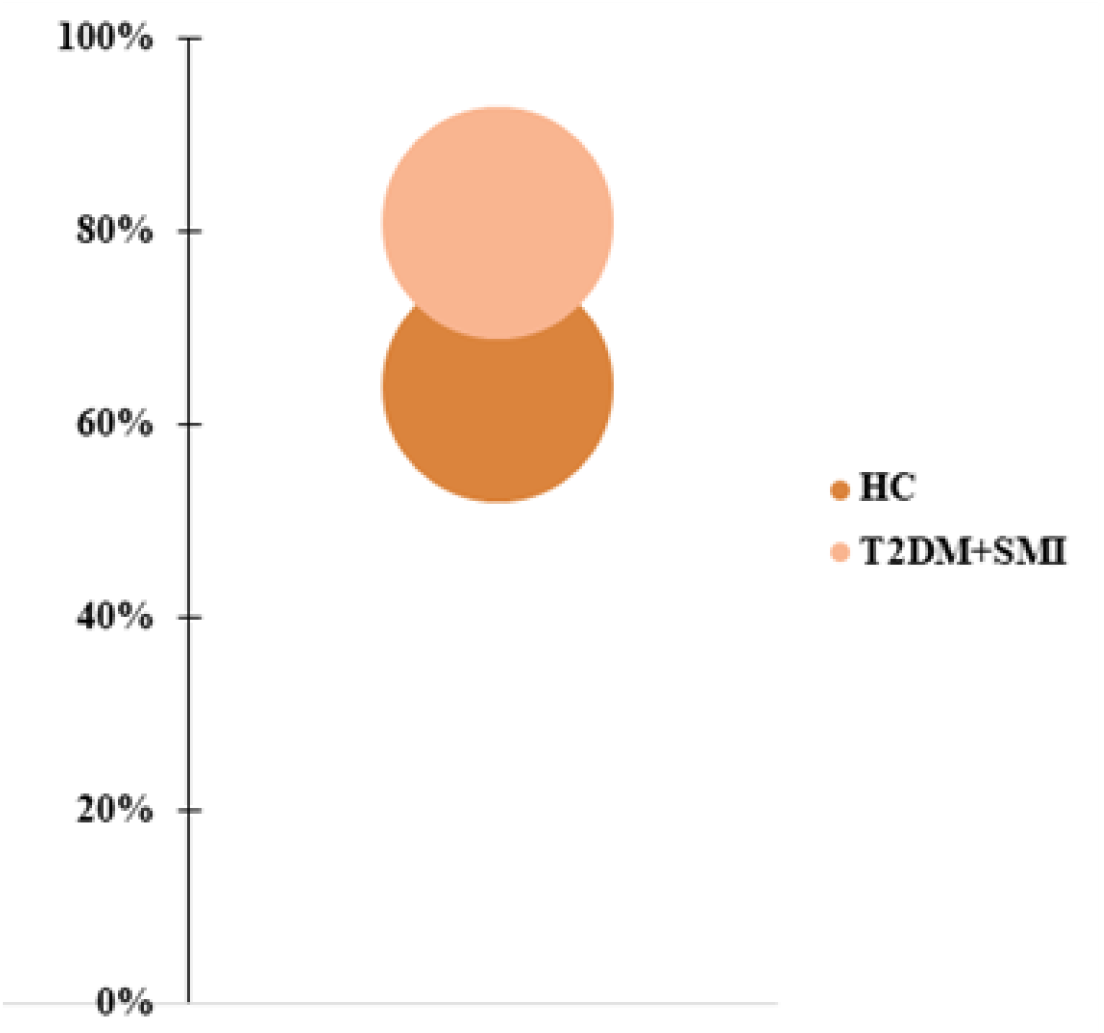
Level of accuracy of transdiagnostic deficits in StM and CR for differentiating individuals with T2DM and SMI from HCs. StM: Short-term memory, CR: Cognitive reserve, T2DM: Type-2 diabetes mellitus, SMI: Severe mental illness, HC: Healthy controls

## DISCUSSION

To the best of our knowledge, this is the first longitudinal study to observe neurocognitive impairment and low social functioning transgressing the nosological boundaries of T2DM, MDD, BD, and SZ. By assessing the performance of these four clinical groups in seven neurocognitive domains, three neurocognitive indices, and three social functioning measures in comparison with that of HCs, we confirmed that the four clinical groups showed several deficits, ranging from moderate-to-large deficits. The most promising transdiagnostic neurocognitive markers consisted of StM and CR. Patients with MDD, BD, and SZ share significant deficits in social functioning. The T2DM group seems to have an intermediate social pattern between HCs and patients with SMI.

The most dramatic general neurocognitive performance deficit was found in individuals with SZ, except in the VM domain, which was more affected in the BD group. Social functioning was most altered in the MDD group. These findings are consistent with previously published literature, which suggests that the neurocognitive alterations present in individuals with SMI and DM2 differ only in their degree of severity [5]. Our findings also coincide with the perspective of the transdiagnostic markers, which show high degree of evidence of neurocognitive comorbidity between SMI and T2DM [37].

These findings reveal potential implications for the care of patients with SMI and T2DM. The specific assessment of neurocognitive performance and social functioning, including quality of life, may facilitate the delimitation of an optimal functional profile to help detect these diseases and prevent them appropriately. For example, a prior identification of underperformance and implementation of measures to correct this may be key elements to a comprehensive therapeutic approach to T2DM individuals. Consequently, assessment protocols should include specific tests for the evaluation of neurocognitive performance and social functioning. This study’s results also indicate the need to generate new treatment protocols focused on the alterations in the neurocognitive processes in these four diagnoses and personalize them to fit the characteristics of the individuals in order to ensure a greater degree of treatment adherence and maximize good therapeutic results. This fact is particularly relevant for patients with T2DM, since neurocognitive and social alterations complicate the control of diabetes and adherence to treatment, thereby negatively affecting prognosis and increasing the risk of diabetes-related complications.[7, 9] Likewise, our findings suggest that StM and CR transdiagnostic neurocognitive deficits should be included when screening individuals for T2DM and SMI. In addition, the degree of neurocognitive impairment in each individual should be assessed. We also highlight the importance of complementary assessments of social functioning when a diagnosis of these diseases is suspected. Thus, we suggest an accessible assessment method for the correct discrimination between individuals with a physical or mental illness and HCs.

This study has a number of limitations to be considered. The initial sample size was small and high experimental mortality was observed after a year of follow-up. Therefore, cross-sectional studies with larger sample sizes could provide more generalizable results. Future studies should also include a more extended follow-up period and greater control of unknown variables to ensure reliable and valid results. Despite these limitations, this is the first known study investigating the possibility of a transdiagnostic overlap in the neurocognitive profiles and social functioning of individuals with SMI and T2DM using a longitudinal design.

Neurocognitive performance and social functioning converge to implicate transdiagnostic disruptions across psychiatric and certain somatic diseases. These findings highlight a common intermediate phenotype, which could help to improve individual responses to treatment. Comprehensive interventions that target cognition improvement could be potentially powerful for ameliorating not only the symptomatic distress but also the lasting functional impairments and poor quality of life. Our findings have an important translational use in terms of identifying common markers for examining the clinical and neurobiological characteristics in these disorders. Nevertheless, future studies should consider the transdiagnostic markers and adopt a longitudinal perspective in order to assess the evolution of neurocognitive performance and social functioning, including quality of life in individuals with SMI and T2DM. In particular, although the performance profile in each clinical group in our study remained relatively stable over time, it can be positively or negatively modified by factors associated with the course of the disease, comorbidities, psychosocial conditions, or certain treatments.[38-40] Therefore, the next step may be to analyze whether neurocognition and other biomarkers predict the real-world functioning in individuals with SMI or T2DM, using a transdiagnostic and longitudinal approach.

## Data Availability

Data are avaliable upon reasonable request writing to

## Acknowledgments

We would like to thank the research participants as well as the staff members of the mental health units of Foios, Catarroja, Paterna, Sagunto, Gandía towns, and the psychiatry outpatient clinic of the University Hospital Dr. Peset and mental health center Miguel Servet, Valencia City.

## REFERENCES

1 Catalá-López F, Gènova-Maleras R, Vieta E, et al. The increasing burden of mental and neurological disorders. Eur Neuropsychopharmacol 2013;23:1337–9.

2 GBD 2019 - Disease and Injury Incidence and Prevalence Collaborators. Global burden of 369 diseases and injuries in 204 countries and territories, 1990-2019: a systematic analysis for the Global Burden of Disease Study 2019. Lancet 2020;396:1204–22.

3 Vancampfort D, Correll CU, Galling B, et al. Diabetes mellitus in patients with schizophrenia, bipolar disorder and major depressive disorder: a systematic review and large-scale meta-analysis. World Psychiatry 2016;15:166–74.

4 Luperdi S, Tabarés-Seisdedos R, Livianos L, et al. Neurocognitive endophenotypes in schizophrenia and bipolar disorder: A systematic review of longitudinal family studies. Schizophr Res 2019;210:21–9.

5 McHutchison CA, Leighton DJ, McIntosh A, et al. Relationship between neuropsychiatric disorders and cognitive and behavioural change in MND. J Neurol Neurosurg Psychiatry 2020;91:245–53.

6 Zhang J, Tian L, Zhang L, et al. Relationship between white matter integrity and posttraumatic cognitive deficits: a systematic review and meta-analysis. J Neurol Neurosurg Psychiatry 2019;90(1):98–107.

7 Karvani M, Simos P, Stavrakaki S, et al. Neurocognitive impairment in type 2 diabetes mellitus. Hormones 2019;18:523–34.

8 Exalto LG, Biessels GJ, Karter AJ, et al. Risk score for prediction of 10 year dementia risk in individuals with type 2 diabetes: a cohort study. Lancet Diabetes Endocrinol 2013;1:183–90.

9 Gilsanz P, Karter AJ, Beeri MS, et al. The bidirectional association between depression and severe hypoglycemic and hyperglycemic events in type 1 diabetes. Diabetes Care 2018;41:446–52.

10 Bora E, Akdede BB, Alptekin K. The relationship between cognitive impairment in schizophrenia and metabolic syndrome: a systematic review and meta-analysis. Psychol Med 2017;47:1030–40.

11 American Psychiatric Association. Manual Diagnóstico y Estadístico de los Trastornos Mentales (DSM 5) Quinta edición. Madrid: Editorial Médica Panamericana 2014.

12 American Diabetes Association. Standards of medical care in diabetes. Diabetes Care 2015;538:1–94.

13 Ramos-Brieva JA, Cordero-Villafáfila AA. Validación de la versión castellana de la Escala de Hamilton para la Depresión. Actas Luso Esp Neurol Psiquiatr 1986;14:324–34.

14 Colom F, Vieta E, Martínez-Arán A, et al. Versión española de una escala de evaluación de la manía: validez y fiabilidad de la Escala de Young. Med Clin 2002;119:366–71.

15 Peralta V, Cuesta MJ. Validación de la escala de síntomas positivos y negativos (PANSS) en una muestra de esquizofrénicos españoles. Actas Luso Esp Neurol Psiquiatr 1994;4:44–50.

16 Vieta E, Torrent C, Martínez-Arán A, et al. Escala sencilla de evaluación del curso del trastorno bipolar: CGI-BP-M. Actas Esp Psiquiatr 2002;30:301–4.

17 Rosa AR, Sánchez-Moreno J, Martinez-Aran A, et al. Validity and reliability of the Functioning Assessment Short Test (FAST) in bipolar disorder. Clin Pract Epidemiol Ment Health 2007;7:3–5.

18 Alonso J, Prieto L, Antó JM. The Spanish version of the SF-36 Health Survey (the SF-36 health questionnaire): an instrument for measuring clinical results. Med Clin 1995;104:771–6.

19 Bobes J, Garcia-Portilla MP, Saiz PA, et al. Quality of life measures in schizophrenia. Eur Psychiatry 2005;20:5313–7.

20 Aliño-Dies M, Sánchez-Ortí JV, Correa-Ghisays P, et al. Grip strength, neurocognition, and social functioning in patients with Type-2 diabetes mellitus, major depressive disorder, bipolar disorder, and schizophrenia. Front Psychol 2020;11:525–31.

21 Correa-Ghisays P, Balanzá-Martínez V, Selva-Vera G, et al. Manual motor speed dysfunction as a neurocognitive endophenotype in euthymic bipolar disorder patients and their healthy relatives. Evidence from a 5-year follow-up study. J Affect Disord 2017;215:156–62.

22 Correa-Ghisays P, Sánchez-Ortí JV, Ayesa-Arriola R, et al. Visual memory dysfunction as a neurocognitive endophenotype in bipolar disorder patients and their unaffected relatives. Evidence from a 5-year follow-up Valencia study. J Affect Disord 2019;257:31–7.

23 San Martín-Valenzuela C, Borras-Barrachina A, Gallego JJ, et al. Motor and cognitive performance in patients with liver cirrhosis with minimal hepatic encephalopathy. J Clin Med 2020;8:21–54.

24 Selva-Vera G, Balanzá-Martínez V, Salazar-Fraile J, et al. The switch from conventional to atypical antipsychotic treatment should not be based exclusively on the presence of cognitive deficits. A pilot study in individuals with schizophrenia. BMC Psychiatry 2010;10:2–10.

25 Benedet MJ, Alejandre MA. TAVEC. Test de Aprendizaje Verbal España-Complutense. Madrid: TEA Ediciones 2014.

26 Golden CJ. Test de Colores y palabras Stroop Manual. Madrid: TEA Ediciones 2001.

27 Grant DA, Berg EA. Test de clasificación de tarjetas Wisconsin Manual. Madrid: TEA Ediciones 2001.

28 Benton AL, Hamsher SK, Sivan AB. Multilingual aplasia examination (2^a^ ed.). Iowa City, IA: AJA Associates 1983.

29 Rosen W. Verbal fluency in aging and dementia. J Clin Neuropsychol 1980;2:135–46.

30 Reitan RM, Wolfson D. The Halstead-Reitan Neuropsychological Test Battery: Theory and Clinical Interpretation. Tucson, Arizona: Neuropsychology Press 1985.

31 Weschler D. Wechsler Adult Intelligence Scale - Third Edition. Escala de Inteligencia Wechsler para Adultos - III. Madrid: TEA Ediciones 1999.

32 Rey A. Test de Copia y de Reproducción de Memoria de Figuras Geométricas Complejas (7^a^ ed.). Madrid: TEA ediciones 1999.

33 Tabarés-Seisdedos R, Salazar J, Selva G, et al. Abnormal motor asymmetry only during bimanual coordination in schizophrenic patients compared to healthy subjects. Schizophr Res 2003;61:245–53.

34 Krull K, Scott J, Sherer M. Estimation of premorbid intelligence from combined performance and demographic variables. J Clin Neuropsychol 1995;9:83–8.

35 Lyman H. Test scores and what they mean. Englewood Cliffs, N.J.: Prentice-Hall 1971.

36 R Core Team. A Language and Environment for Statistical Computing. R Foundation for Statistical Computing, Vienna, Austria 2016. URL: https://www.R-project.org/.

37 Tabarés-Seisdedos R, Baudot A. Direct and inverse comorbidities between complex disorders. Front Physiol 2016;7:1–2.

38 Bonnin CM, Torrent C, Arango C, et al. Functional remediation in bipolar disorder: 1-year follow-up of neurocognitive and functional outcome. Br J Psychiatry 2016;208:87–93.

39 Sánchez-Valle J, Tejero H, Fernández JM, et al. Interpreting molecular similarity between patients as a determinant of disease comorbidity relationships. Nat Commun 2020;11:28–54.

40 Tabarés-Seisdedos R, Rubenstein JL. Inverse cancer comorbidity: a serendipitous opportunity to gain insight into CNS disorders. Nat Rev Neurosci 2013;14:293–304.

